# Prevalence of malformations of cortical development in patients with suspected epilepsy based on a clinical MRI dataset

**DOI:** 10.64898/2026.08.19.26360591

**Authors:** Llucia Coll, Júlia Díaz-i-Calvete, Alice Schiavone, Helene Kaas, Martin Prener, Vincent Beliveau, Gitte Moos Knudsen, Lars Hageman Pinborg, Melanie Ganz

**Affiliations:** Neurobiology Research Unit, Copenhagen University Hospital, Copenhagen, Denmark; Danish Research Centre for Magnetic Resonance, Centre for Functional and Diagnostic Imaging and Research, Copenhagen University Hospital - Amager and Hvidovre, Copenhagen, Denmark; Department of Computer Science, University of Copenhagen, Copenhagen, Denmark; Epilepsy Clinic, Department of Neurology, Copenhagen University Hospital – Rigshospitalet, Copenhagen, Denmark; Institute for Human Genetics, Medical University of Innsbruck, Innsbruck, Austria; Institute of Clinical Medicine, University of Copenhagen, Copenhagen, Denmark

## Abstract

**Objective:** To estimate the prevalence of epilepsy-associated malformations of cortical development (MCDs) in Eastern Denmark, and to validate whether epilepsy prevalence in the same population is consistent with national estimates.

**Methods:** A retrospective cohort study of people registered with ICD-10 code DG40* and/or DZ033A from 1998 up to 1 July 2023 was conducted. The study population was defined as all living residents in Eastern Denmark with at least one recorded hospital-patient contact within the year preceding 1 July 2023. Magnetic resonance imaging (MRI) availability was required to assess presence of any MCD. MRI radiology reports were manually reviewed or evaluated using a language model to identify MCDs, including encephalocele, focal cortical dysplasia (FCD), hemimegalencephaly, heterotopia, hypothalamic hamartoma, lissencephaly, polymicrogyria and schizencephaly. Prevalence estimates were calculated for each MCD subtype and for epilepsy overall, and compared with the available literature.

**Results:** On 1 July 2023, 28,739 people met inclusion criteria, and 14,434 had an available brain MRI, including radiological description of possible MCDs. The prevalence per 100,000 population was 1044.6 (95% CI 1032.6 to 1056.6) for epilepsy and 32.1 (95% CI 30.1 to 34.3) for any MCD associated with seizures. Reported MCD prevalence in the literature, when existent, was derived from pediatric age-ranged selected cohorts, except for FCD. No prevalence estimates for hemimegalencephaly and heterotopia were identified.

**Significance:** We presented the first population-based estimates of seizure-associated MCD prevalence in a large all-age cohort. Direct comparison with prior literature was prevented due to differences in study design and population structure, but epilepsy prevalence was consistent with previously reported national estimates.

**Key points:**

- First prevalence estimates of malformation of cortical development presenting with seizures on a large all-age cohort.
- Epilepsy prevalence estimates align with Denmark’s nationwide estimates previously reported.
- Language models used on nation-wide registries can contribute to elucidate the epidemiology of rare conditions.

## 1 Introduction

Epilepsy is amongst the most common serious neurological disorders, affecting around 50 million individuals of all ages worldwide [1]. It is characterised by abnormal synchronous neuronal activity that leads to recurrent, unprovoked seizures [2].Its associated risk of premature death, together with its stigma and increasing burden-economic, psychosocial, physical, and mental-makes epilepsy a public health imperative target [1, 3, 4]. To enable evidence-based actions that improve the care and treatment of patients, accurate and up-to-date data on epilepsy epidemiology are crucial [1]. A recent systematic analysis monitored the epilepsy burden at global, regional, and national levels from 1990 to 2021 [5]. In this and similar burden analyses, epilepsy estimates distinguish between primary (idiopathic) and secondary epilepsies [6, 5]. However, these categories differ from the current International League Against Epilepsy (ILAE) framework, which classifies epilepsy considering several dimensions including aetiology [7]. Structural aetiologies include malformations of cortical development (MCDs), which are a clinically relevant cause of epilepsy, but their prevalence at the population level remains insufficiently characterised. Building on the need to refine broad epidemiological estimates into more specific aetiological data, the present study aims to characterise the prevalence of MCDs associated with epilepsy, addressing a current gap with direct relevance to diagnostic evaluation.

Unlike generalised seizures, focal seizures originate within networks limited to one hemisphere [7, 8]. These seizure types predominate across all ages, and are correlated with a higher risk of seizure recurrence [9, 4]. Importantly, focal epilepsy is a significant predictor of drug-resistant epilepsy (DRE), where seizure-free status is not achieved despite anti-epileptic medications [10, 11]. In DRE, surgical treatment may be considered, and the identification of a structural aetiology can support the presurgical evaluation [7, 12].

MCDs are considered a heterogeneous family of congenital brain malformations originating from disturbed development of the cerebral cortex in fetal life [13, 14]. They can be challenging to identify, often requiring electro-clinical profiling to guide epilepsy-specific magnetic resonance imaging (MRI) protocols and experienced neuroradiologist assessments [15, 7]. Consequently, studies that consistently estimate the incidence and prevalence of specific MRI-detected abnormalities, particularly MCDs in epilepsy, remain scarce. At the same time, the growing volume of clinical data stored in electronic health records (EHRs) and radiology databases represents both a data-management challenge and an opportunity for epidemiological research [16].]. Manual review of free-text clinical notes to identify and count patients with a given condition is resource-demanding, and difficult to scale. Natural language processing (NLP) and, more recently, large language models have emerged as powerful tools to address this bottleneck [17]. In radiology, NLP-based methods have been used to automatically extract neurological findings from brain imaging reports, enabling large-scale phenotyping and pathology prevalence estimation without the resource demands of manual text review [18]. Building on this, Schiavone *et al.* [19] recently proposed a language model (LM) for processing radiology report from multiple imaging modalities, with epilepsy classification among its evaluated tasks.

Assessing the frequency of different MCD subtypes in patients with epilepsy can improve our understanding of the distribution of these abnormalities and provide a reference for future epidemiological, clinical, and methodological studies. The aim of this study is to estimate the prevalence of seizure-associated MCDs in Eastern Denmark, both among persons with suspected epilepsy, and in the general population. We included the most relevant MCDs recognised as structural causes of epilepsy by the ILAE: encephalocele, focal cortical dysplasia (FCD), hemimegalencephaly, heterotopia, hypothalamic hamartoma, lissencephaly, polymicrogyria (PMG) and schizencephaly [7].

### Overview of cortical malformations in epilepsy

The epidemiology of MCDs is highly based on classification of resected tissue samples from epilepsy surgeries [12]. MCDs are responsible for up to 40% of DRE and can cause severe cognitive impairments [20]. In 9523 DRE patients who underwent epilepsy surgery, 19.8% were categorised as MCDs, being also the case in 39.3% of samples obtained from children [21]. A recent review reported an overall prevalence of MCDs of 6.52 ± 1.89 in 100,000 children, slightly lower in adults (see Table 2) [12]. Below, we review the existing literature on published cohorts that report epidemiological estimates for MCD subtypes (more details in the Supplementary material). It is worth considering that MCDs can present in seizure-free individuals. Furthermore, the accurate prevalence of most specific structural abnormalities is not available from large homogeneous datasets. Our work focuses on MCDs specifically in patients with suspected epilepsy on a region-wide level, so the comparison with the existent literature is limited (see Discussion).

#### Focal cortical dysplasia

FCDs are localised regions of malformed cerebral cortex presenting cortical disorganisation, large abnormal neurons, and, in half of the patients, balloon cells. These lesions can lead to cognitive delays and early onset of seizures, with varying clinical implications according to subtype [22].FCD has previously been reported as the most common type of MCD in patients undergoing epilepsy surgery, accounting for 70.6% of these cases, of which children was the largest subgroup (52.7%) [21]. In adults, the prevalence of FCD was 4.24 per 100,000 people, and 4.43 for children [12].

#### Polymicrogyria

PMG is characterised by abnormally small misfolded gyri that results from disturbances in neuronal migration [14]. PMG is estimated to constitute 16% of MCDs [23]. The prevalence of PMG is estimated at 2.3 per 10,000 children, with an annual incidence of 1.9 per 10,000 person-year. Epilepsy was diagnosed in 54% of cases, with most cases being DRE [24].

#### Schizencephaly

Schizencephaly is defined by clefts extending through the hemispheres from the ventricles to the pial surface. It is estimated to occur in 0.54 - 1.54 per 100,000 births, based on data from the United States, European countries, and Japan [25, 26, 27, 28].

#### Heterotopia

Grey matter heterotopia (GMH) is characterised by neurons at abnormal positions. In subependymal heterotopia, the disorder presents clusters of neurons along the lateral ventricle walls; in focal subcortical heterotopia, nodules are formed in the white matter, and in subcortical band heterotopia (SBH), bands of subcortical heterotopic neurons form between the lateral ventricle and the cerebral cortex bilaterally, predominantly seen in females [14, 29]. SBH is considered rare, [29], while GMH appears to be more common than was once thought. GMH may constitute about 15% of MCDs and can be found in about 2% of patients with epilepsy, often DRE. Its general-population prevalence is unknown, and its presence in asymptomatic people remains debated [30, 31].

#### Lissencephaly

Lissencephaly refers to a reduction in sulcation, meaning the surface of the brain appears smooth. The prevalence is estimated at around 0.4 - 1.2 per 100,000 births for type I [32]. This type is associated with SBH [33].

#### Hemimegalencephaly

Hemimegalencephaly is characterised by abnormal enlargement of one cerebral hemisphere, affecting all of it or only individual lobes [34]. It is often associated with DRE, and it frequently presents in early infancy [35]. It is estimated that there are 1 - 3 cases of hemimegalencephaly per every 1,000 children with epilepsy, and it seems to represent up to 14% of cases of MCDs [34].

#### Hypothalamic hamartoma

Hypothalamic hamartomas are non-progressive lesions that may be intrahypothalamic or parahypothalamic. Both types are associated with a disabling clinical course associated, multiple seizure types, cognitive decline and psychiatric symptoms [36]. It is a rare condition estimated to occur in about 1.5 in 100,000 children [37], slightly predominantly among males.

#### Encephalocele

Encephaloceles are rare neural tube defects leading to herniations of the brain parenchyma through the dura matter and skull [38, 39]. The incidence of congenital encephalocele is estimated at 1 in 10,000 live births, although this rate may be in fact higher, given the pregnancy terminations that follow many prenatal diagnoses [39].

## 2 Materials and methods

### Study design

This population-based cohort study is based on all persons with epilepsy and suspected epilepsy in the *Sundhedsplatform* (SP), a Danish electronic health system, living in Region Zealand and Capital Region of Denmark (Eastern Denmark) from 1998 up to 1 July 2023. Due to a change in the electronic health system in 2016, most analysed data is from the period 2016 to 2023.

Ethical approval for data collection was granted by the relevant ethics committees (CVK-2112460) and approved for data processing by the Danish Data Protection Authorities. According to Danish law, informed consent is not required for this type of retrospective study. All data were de-identified and data processing undertaken on a GDPR-compliant system at the Neurobiology Research Unit, Copenhagen University Hospital.

### 2.1 Population and cohorts

The entire living population in Eastern Denmark was used as denominator for prevalence calculations. Demographics of this regional background population were obtained from Statistics Denmark (www.dst.dk) for the relevant Regions, sexes and age groups as of July 2023.

All prevalence estimates calculated here were restricted to persons coded with ICD-10 codes DG40* and/or DZ033A in the EHR. Demographic data for the entire coded cohort were obtained through their CPR numbers. The CPR is a 10-digit personal identification number unique to each Danish resident, used as patient identifier within the healthcare system. It encodes date of birth and sex.

#### Definition of analytical cohort

The SP registers the complete clinical record for all patients registered in Eastern Denmark health system. However, deceased persons are not systematically removed, nor individuals relocated from the country or Region, transferred to private healthcare, or with discontinued follow-up for other reasons, including no final epilepsy diagnosis at discharge.

To account for these scenarios, eligibility was assessed at the extraction date (1 July 2023) through screening steps based on EHR data. First, we searched for the term ”mors-notat”, commonly used in clinical documentation to indicate death on the EHR. After excluding documented deaths, we further restricted the sample to patients with recent entries in their clinical records. The latest date was assumed to reflect the last recorded patient-hospital contact, regardless of whether it was epilepsy-related. The final analytical cohort was defined by requiring at least one recorded contact within the year preceding 1 July 2023. This aimed to ensure that our coded suspected epilepsy cohort (CSEC) were living in Eastern Denmark.

#### Sub-group with available MRI

The cohort used to evaluate the different MCDs was defined as the CSEC with available MRI data obtained from data in the PACS system. For each MRI acquisition, the associated radiology report was used to classify radiological findings corresponding to each specific MCD. Label extraction is described in the following section.

### MCD identification from reports

To identify patients with MCDs, we extracted relevant findings from MRI radiological reports using MOSAIC-12B, a LM trained on chest X-ray radiological reports in English, Spanish, and French [19]. As LMs may exhibit prompt sensitivity and hallucinations, model outputs were validated against manually annotated reports [40]. Performance was assessed using the F1 score. More details on model deployment are provided in the Supplementary Material. To estimate MCD prevalences, we considered the three most recent radiological reports per patient and classified a patient as positive if at least one report was positive.

### Statistical analyses

The prevalence of suspected epilepsy (*P_CSEC_*) was calculated as the number of persons meeting the inclusion criteria (*N_CSEC_*) divided by the total population living in Eastern Denmark on 1 July 2023 (*N*). The inclusion criteria were the presence of the aforementioned ICD-10 codes and with at least one patient-hospital contact during the year prior to 1 July 2023. Estimates were stratified by sex (female, male) and age group (10-year intervals).

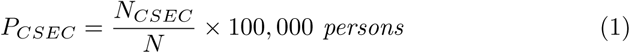

The prevalence of CSEC with available MRI examination (*P_CSEC^MRI_*) was defined as the number of persons with suspected epilepsy who underwent at least one brain MRI during the study period (*N_CSEC^MRI_*) divided by the total population (*N*). It should be noted that *P_CSEC^MRI_* is a reference estimate of the cohort with sufficient data to derive the prevalence of MCDs. We assume that any MRI were taken as part of a standard epilepsy diagnostic workup within the CSEC population.

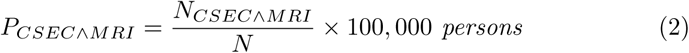

The prevalence of MCDs among persons with suspected epilepsy within the population was calculated as the number of persons in CSEC with an identified MCD from MRI radiology reports, in relation to the total population (*N*). Estimates are provided for all MCDs combined and per subtype (*k*: encephalocele, FCD, hemimegalencephaly, heterotopia, hypothalamic hamartoma, lissencephaly, PMG and schizencephaly). Given that MRI was not available for all CSEC individuals, 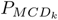 may represent lower-bound estimates, as individuals with suspected epilepsy and underlying MCD who did not undergo MRI were not included in *N_CSEC^MRI_*.

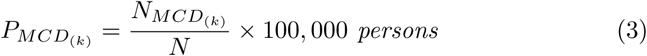

All prevalence rates were expressed per 100,000 persons to align with standard reporting in the literature and facilitate comparison. 95% confidence intervals (CIs) were estimated using the Wilson method.

Sensitivity analyses were performed using alternative inclusion criteria: (i) only excluding persons with confirmed death record; and (ii) requiring active 3- and 5-year patient-hospital contact prior to 1 July 2023.

The population-based prevalence estimates of suspected epilepsy were indirectly compared to estimates reported on the whole Danish population, including Christensen *et al.* [41], as well as the 2023 estimates from the Global Burden of Disease Study (GBD) reported for Denmark [42]. MCD prevalence estimates in patients with epilepsy were compared with published epidemiology specific to each type of cortical malformations, when available.

## 3 Results

### Population

On 1 July 2023, a total of 2,751,313 individuals (1,394,079 [50.7%] female and 1,357,234 [49.3%] male) were living in Eastern Denmark. On the same date, 43,145 people (21,017 [48.7%] female and 22,128 [51.3%] male) had received an ICD-10 code DG40* and/or DZ033A at any time and were registered in the system. Based on EHR data, 2,877 people were confirmed deceased and therefore excluded from the study. We also excluded 56 individuals with unavailable EHR or MRI data. The remaining 40,212 (19,810 [49.3%] female and 20,402 [50.7%] male) were further screened for a minimum of 1-year recorded hospital-patient contact prior to the study end date (July 2023). This resulted in 28,739 (14,347 [49.9%] female and 14,392 [50.1%] male) presumably living persons with suspected epilepsy. The full inclusion and exclusion criteria are summarised in Figure 1. Regarding brain MRI availability, 14,434 individuals (7,300 [50.6%] female and 7,134 [49.4%] male) had at least one brain MRI. Age-range distribution of the cohort with and without MRI availability is represented in Figure 1.

**Figure 1:**
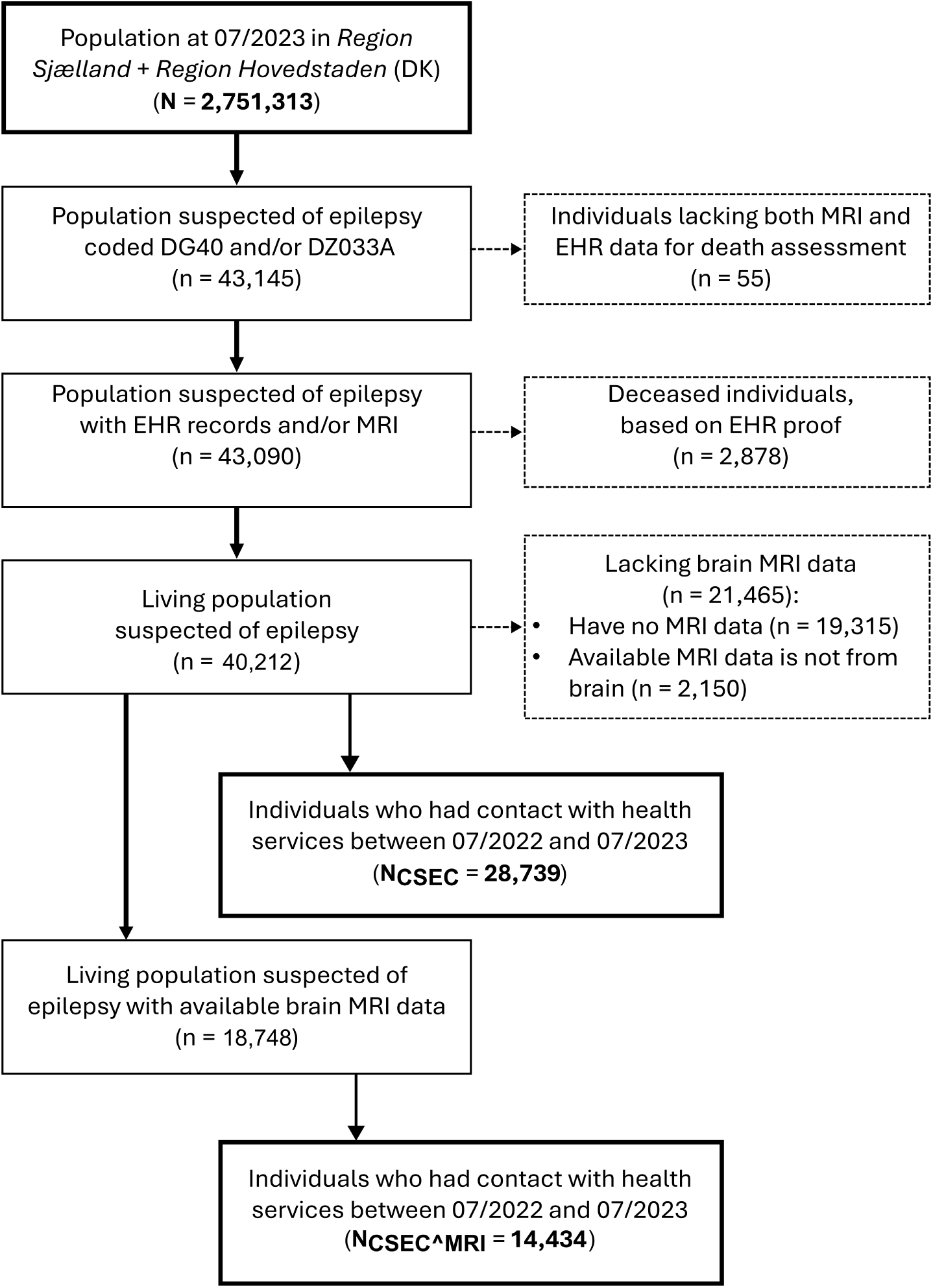
Flowchart illustrating the inclusion (solid lines/boxes) and exclusion (dashed lines/boxes) processes of the individuals in the population. The final study sample used for analysis is highlighted in bold. *N* = total population; *n* = sample; *N_CSEC_* = number of individuals of the coded suspected epilepsy cohort; *N_CSEC^MRI_* = number of individuals of the coded suspected epilepsy cohort who underwent MRI.

Regarding brain MRI availability, 14,434 individuals (7,300 [50.6%] female and 7,134 [49.4%] male) had at least one brain MRI. Age-range distribution of the cohort with and without MRI availability is represented in Figure 2.

**Figure 2:**
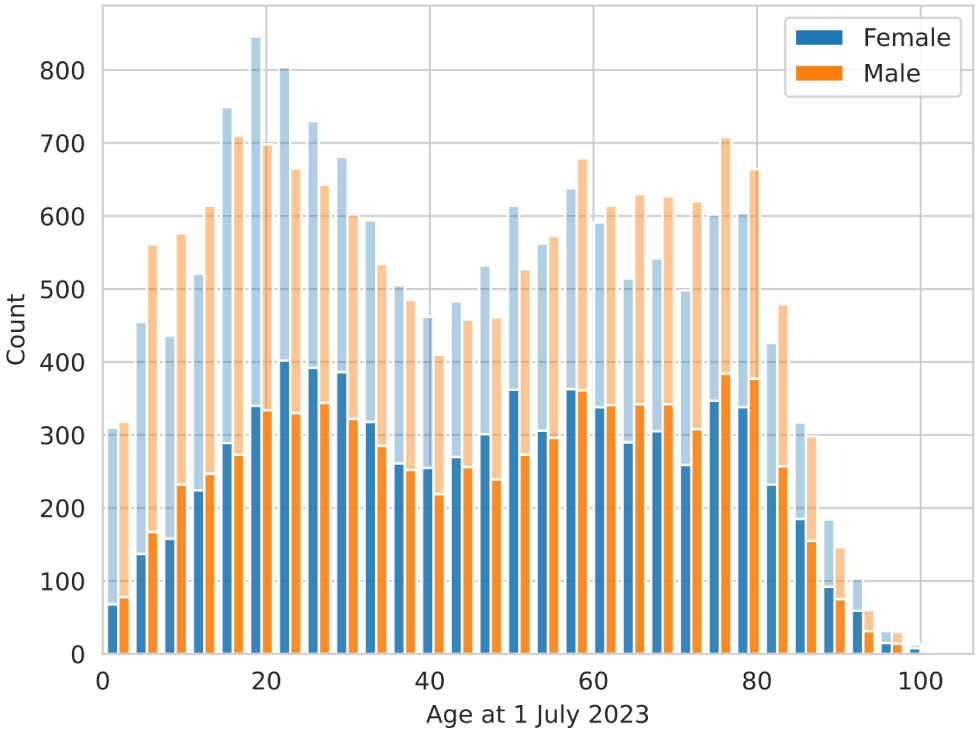
Age at 1 July 2023 and sex distributions across the cohort of patients with suspected epilepsy. MRI availability is also distinguished according to the bar lightness (dark: MRI, light: no-MRI).

### Language model performance

Performance comparison between the LM vs manual annotations is shown in the Supplementary Table S1. With the zero-shot setting, the LM achieved high specificity, but low precision and recall on average. When fine-tuned for the task, the model performance improved, from a mean 0.58 to 0.73 F1. Excluding lissencephaly and schizencephaly, which both had a single data point in the test set, the mean was 0.80 F1. The MCD with the largest improvement was hypothalamic hamartoma (from 0.36 to 0.76 F1). For FCD, the most present MCD in our dataset, fine-tuning helped also on the other classes in our taxonomy. We adjusted our final prevalence estimates by manually annotating reports based on the automatically identified MCDs. However, FCD (0.80 F1) and heterotopia (0.81 F1) were excluded from this manual review due to their high sample volume, which rendered individual verification unfeasible.

### Prevalence estimates

#### Suspected epilepsy and MRI

The *P_CSEC_* on 1 July 2023 was 1044.6 (95% CI 1032.6 to 1056.6) per 100,000 population (1029.1 [95% CI 1012.5 to 1046.0] per 100,000 female population and 1060.4 [95% CI 1043.3 to 1077.8] per 100,000 male population). The *P_CSEC^MRI_* was 524.6 (95% CI 516.2 to 533.2) per 100,000 population (523.6 [95% CI 511.8 to 535.8] per 100,000 female population and 525.6 [95% CI 513.6 to 537.9] per 100,000 male population). Age-specific estimates of suspected epilepsy and suspected epilepsy with MRI examination are shown in Figure 3. Sensitivity analyses with alternative inclusion criteria used for prevalence estimates are summarised in the Supplementary Table S2 (see Supplementary Figure S1 for age-specific distributions).

**Figure 3:**
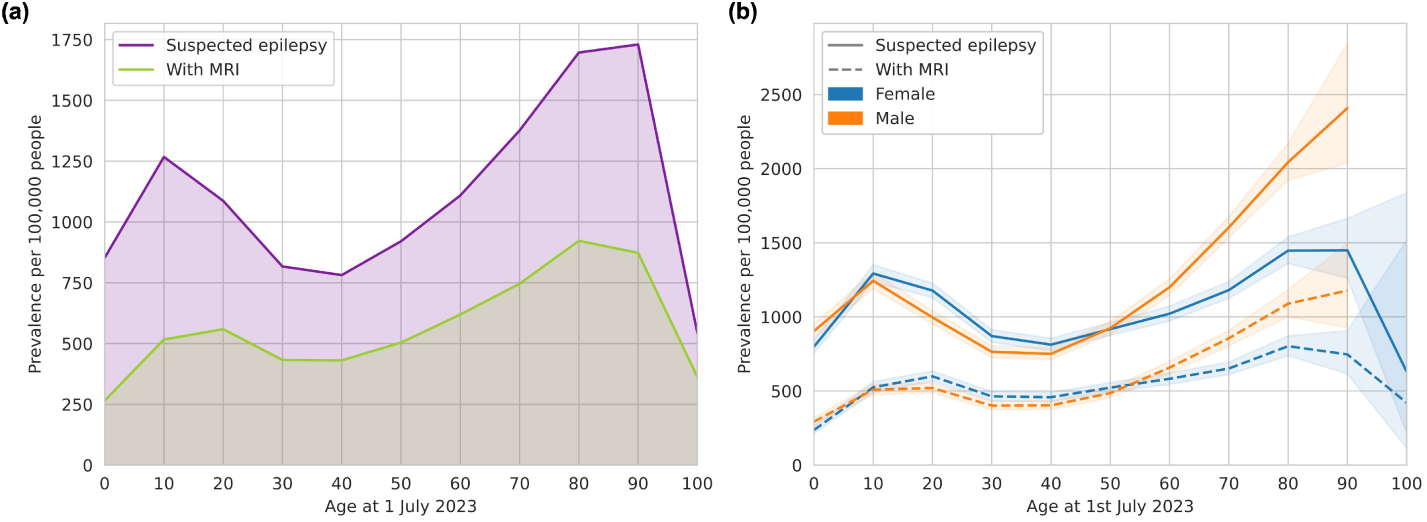
Age-specific prevalence of (i) suspected epilepsy and (ii) those who underwent brain MRI acquisition as part of their clinical examination for epilepsy in Eastern Denmark on 1 July 2023: (a) overall population and (b) stratified by sex.

#### MCD

The prevalence of presenting any MCD (*P_MCD_*) was 31.3 (95% CI 29.3 to 33.5) per 100,000 population (30.8 [95% CI 28.0 to 33.8] per 100,000 female population and 31.9 [95% CI 29.0 to 35.1] per 100,000 male population). Table 1 shows the prevalence for each single MCD. The age-specific MCD prevalence estimates are shown in Figure 4. Sensitivity analyses results are provided in the Supplementary Table S3.

**Figure 4:**
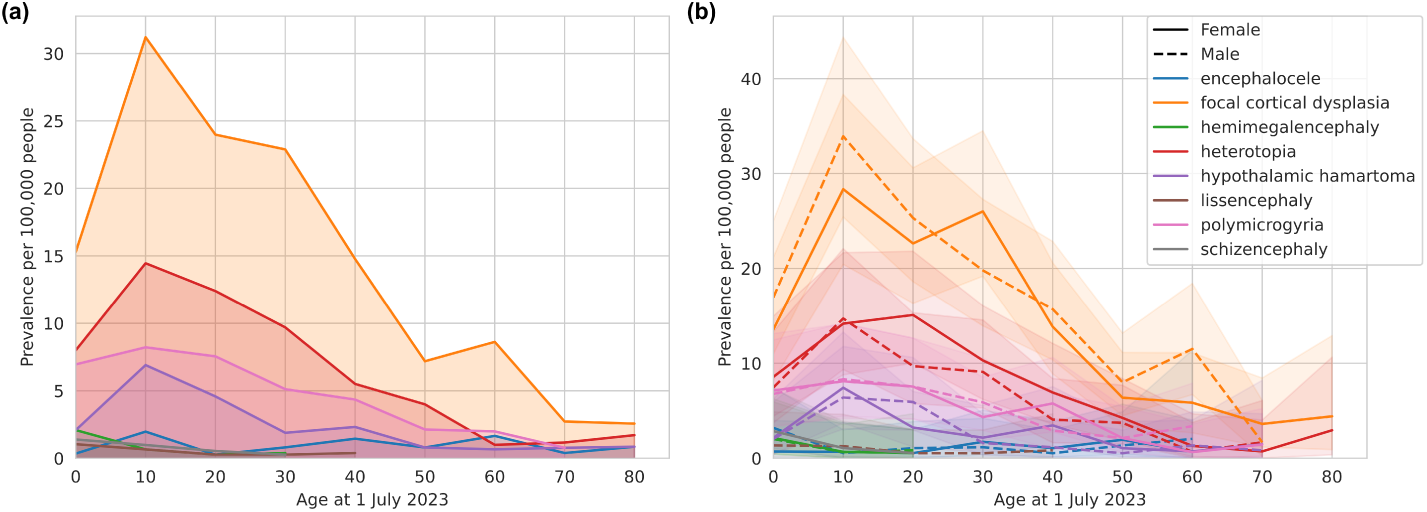
Age-specific prevalence of malformations of cortical development (MCDs) associated with suspected epilepsy in Eastern Denmark on 1 July 2023: (a) overall population and (b) stratified by sex

**Table 1:** Prevalence of (i) suspected epilepsy presenting any cortical malformation and (ii) suspected epilepsy with a specific cortical malformation (encephalocele, focal cortical dysplasia, hemimegalencephaly, heterotopia, hypothalamic hamartoma, lissencephaly, polymicrogyria, schizencephaly), against general population in Eastern Denmark on 1 July 2023, stratified by sex. MCD: malformation of cortical development, FCD: focal cortical dysplasia, PMG: polymicrogyria.

|  | Overall |  | Female |  | Male |  |
| --- | --- | --- | --- | --- | --- | --- |
|  | N | Prevalence (95% CI) per 100,000 | N | Prevalence (95% CI) per 100,000 | N | Prevalence (95% CI) per 100,000 |
| Any MCD | 862 | 31.33 (29.31 to 33.49) | 429 | 30.77 (28.0 to 33.83) | 433 | 31.90 (29.04 to 35.05) |
| Encephalocele | 26 | 0.95 (0.65 to 1.38) | 12 | 0.86 (0.49 to 1.50) | 14 | 1.03 (0.61 to 1.73) |
| FCD | 427 | 15.51 (14.12 to 17.06) | 204 | 14.63 (12.76 to 16.78) | 223 | 16.43 (14.41 to 18.73) |
| Hemimegalencephaly | 10 | 0.36 (0.20 to 0.67) | 5 | 0.36 (0.15 to 0.84) | 5 | 0.37 (0.16 to 0.86) |
| Heterotopia | 191 | 6.94 (6.02 to 8.00) | 105 | 7.53 (6.22 to 9.12) | 86 | 6.34 (5.13 to 7.82) |
| Hypothalamic hamartoma | 67 | 2.44 (1.92 to 3.09) | 34 | 2.44 (1.75 to 3.41) | 33 | 2.43 (1.73 to 3.41) |
| Lissencephaly | 8 | 0.29 (0.15 to 0.57) | 1 | 0.07 (0.01 to 0.41) | 7 | 0.52 (0.25 to 1.06) |
| PMG | 123 | 4.47 (3.75 to 5.33) | 61 | 4.38 (3.41 to 5.62) | 62 | 4.57 (3.56 to 5.86) |
| Schizencephaly | 10 | 0.36 (0.20 to 0.67) | 7 | 0.50 (0.24 to 1.04) | 3 | 0.22 (0.08 to 0.65) |

### Literature comparison

Table 2 shows a summarised overview of published studies on MCD cohorts, alongside our corresponding estimates calculated using the same age-range inclusion criteria. Notably, no prevalence estimates for hemimegalencephaly and heterotopia were identified in the literature.

**Table 2:** Prevalence of (i) any cortical malformation and (ii) single cortical malformations (encephalocele, focal cortical dysplasia, hemimegalencephaly, heterotopia, hypothalamic hamartoma, lissencephaly, polymicrogyria, schizencephaly) by specific age ranges available in current literature, with corresponding estimates from our cohort with suspected epilepsy, calculated under matched inclusion criteria. MCD: malformation of cortical development, FCD: focal cortical dysplasia, PMG: polymicrogyria, ’-’: non-available.

|  | Age range (years) | Literature |  |  | Our cohort |  |
| --- | --- | --- | --- | --- | --- | --- |
|  |  | N | Prevalence (95% CI) per 100,000 | Reference | N | Prevalence (95% CI) per 100,000 |
| Any MCD | < 18 | - | 6.52 SD = 1.89 | [12] | 260 | 49.05 (43.44 to 55.39) |
|  | > 18 | - | 6.03 SD = 0.67 |  | 602 | 27.10 (25.02 to 29.35) |
| Encephalocele | < 2 | 532 | 10.30 (9.40 to 11.20) | [43] | - | - |
|  | < 5 | 40 | 10.70 (3.40 to 18.10) | [44] | 1 | 0.69 (0.12 to 3.90) |
| FCD | < 18 | - | 4.43 SD = 0.90 | [12] | 118 | 22.26 (18.59 to 26.66) |
|  | > 18 | - | 4.24 SD = 0.34 |  | 309 | 13.91 (12.44 to 15.55) |
| Hemimegalencephaly | - | - | - | [34] | - | - |
| Heterotopia | - | - | - | [29] | - | - |
| Hypothalamic Hamartoma | - | - | 1.5 | [37] | - | - |
| Lissencephaly | < 10 | 22 | 1.17 | [45] | 3 | 1.04 (0.35 to 3.06) |
| PMG | < 18 | 109 | 23.10 (19.00 to 28.00) | [24] | 38 | 7.17 (5.22 to 9.84) |
| Schizencephaly | < 1 | 63 | 1.54 | [26] | - | - |
|  | < 5 | 38 | 1.48 (1.01 to 1.95) | [25] | - | - |
|  | < 7 | 10 | 5.40 (2.00 to 8.70) | [27] | 1 | 0.49 (0.09 to 2.78) |

## 4 Discussion

In this population-based study of almost 3 million people living in Eastern Denmark, we estimated the prevalence of suspected epilepsy and MCDs, using routinely collected data and advanced language models techniques. Our prevalence estimates for epilepsy are consistent with previously reported numbers [41, 42]. In contrast, the available literature on MCDs is considerably more limited and heterogeneous, with substantial variation in study design, case definitions, and populations. Consequently, interpretation is restricted to comparisons with individual studies and their specific cohort definition.

Our prevalence estimates of suspected epilepsy are comparable to those reported by Christensen *et al.* [41], where the prevalence of confirmed epilepsy between 2009 and 2018 was estimated at 697 (95% CI 691 to 704) per 100,000 population. This is lower than our overall prevalence of 1044.6 (95% CI 1032.6 to 1056.6) per 100,000 population. However, when broader case definitions (previous diagnosis and anti-seizure medication prescriptions) were applied, prevalence increased to 846 (95% CI 839 to 854) and 1218 (95% CI 1209 to 1227) per 100,000 population, respectively[41]. These estimates are closer to ours, potentially reflecting the broader inclusion criteria used to identify suspected epilepsy, rather than confirmed cases. Further insight can be gained with our population prevalence of suspected epilepsy with MRI examination, which was 524.6 (95% CI 516.2 to 533.2) per 100,000 population. Compared with their main definition based on confirmed diagnosis, our MRI-based estimate likely captures a clinically selected population with suspected epilepsy who underwent diagnostic evaluation, which may contribute to the comparability of the estimates. Furthermore, our prevalence without follow-up restrictions (see Supplementary Table S2) was within the range of estimates reported by Christensen *et al.* primary definition [41]. Discrepancies in inclusion criteria add to differences in study population characteristics, as Christensen *et al.* included nationwide Danish data, whereas our study focused on two highly populated regions (Zealand and Capital Region). This pattern is also observed in comparison with estimates from the GBD 2023 study, which reported a global prevalence of 683.4 (95% CI 587.6 to 787.4) per 100,000 population and a prevalence of 522.4 (95% CI 326.6 to 705.9) per 100,000 population in Denmark [42].

Although epilepsy epidemiology is well-established in pediatric and adult populations, the frequency of many MCDs, particularly non-FCD, remains poorly characterised. Consequently, comparisons with the literature are less straight-forward. For many lesion types, epidemiological estimates remain unknown [12], and the limited number of reported cases constrains the identification of robust demographic patterns [46].

A key difference from our study compared to the literature is the cohort composition and the association of MCDs to epilepsy symptoms. Most studies focus exclusively on infant populations, often within narrowly defined age ranges [25, 26, 27, 44, 45]. Restricting our estimates to those age-ranges reported in the literature substantially altered our results, observing no clear agreements. The reported estimates of FCD, for example, (4.43 [SD=0.9] and 4.24 [SD=0.34]) are much lower than ours (22.26 [95% CI 18.59 to 26.66] and 13.91 [95% CI 12.44 to 15.55]). However, neither the underlying population used to calculate prevalence nor the number of positive cases were reported by López-Rivera *et al.* [12]. A larger population at risk or under-identification of cases could explain their lower reported prevalence. In contrast, for PMG, the literature prevalence for people younger than 18 is higher (23.10 [95% CI 19 to 28]) than our estimate (7.17 [95% CI 5.22 to 9.84]) [24]. In this case, they studied the presence of pediatric PMG, but the associated clinical symptomatology was not always necessarily seizures: only 54% of children diagnosed with PMG presented epilepsy. This is therefore not directly comparable to our analysis, where we specifically assess the prevalence of PMG associated to epilepsy, and explains our lower estimates, also when calculating our all-age PMG prevalence (4.47 [95% CI 3.75 to 5.33]), which also lack correspondence in the literature.

For some MCDs, such as hemimegalencephaly [34] and hypothalamic hamartoma [37], the prevalence values reported are estimates drawn from case-studies or earlier literature with limited methodological transparency and unclear underlying assumptions. For others, including heterotopia, prevalence remains uncertain [29].

Differences in reported prevalence estimates largely reflect heterogeneity in surveillance methodologies, including variation in case definitions, ascertainment, and follow-up [47]. The inclusion of live births only, stillbirths, late fetal losses and terminations of pregnancy-in some cases, due to prenatal MCD diagnoses-can also impact the results [25, 43]. In addition, variation in clinical severity may further influence these estimates. Some abnormalities are associated with severe life-threatening presentations, affecting life expectancy and thereby their representation in adult populations [48]. This highlights the importance of assessing pediatric and adult populations separately. Concurrently, lesions with milder or asymptomatic presentations are less likely to be consistently identified and reported, potentially leading to an underestimation of their true prevalence [43].

Our methodology is not without limitations. Patient identification from the EHR was solely based on ICD-10 codes, which have resulted in a broader inclusion, as registry-based epilepsy diagnoses alone do not always reflect confirmed epilepsy [49]. EHR data are dependent on the health system and internal structure which may vary in their degree of standardisation. In this study, we primarily relied on the use of radiology reports for the identification of different MCDs, and EHR for complementary case information. Although these components provide valuable information, they are subject to variability in reporting quality and completeness. Furthermore, LM-based identification of MCDs, particularly FCD and heterotopia, was not fully manually validated and is therefore subject to diagnostic uncertainty. MRI availability was also unlikely to be random, as imaging is influenced by clinical indication. Consequently, the MRI subgroup may not be representative of all patients with suspected epilepsy, which should be considered when interpreting MCD prevalence estimates. Loss to follow-up may reflect death, migration from the region, or treatment transfer to the private sector, resulting in absence of subsequent record in the public health system. The selected 1-year follow-up threshold was evaluated against alternative windows of 3 and 5 years, as well as without follow-up restrictions (see Supplementary Material). Across these scenarios, the age-specific distribution of prevalence remained consistent, with differences only observed in absolute counts and, consequently, in prevalence estimates.

Given the relatively low prevalence of some MCDs, natural regional variations are expected to appear, especially in areas with a smaller number of births (i.e., *<* 100, 000). Populations that differ in age, ethnicity, access to obstetric care and other regional traits may also differ in underlying susceptibilities and/or risk factors that could impact the birth prevalence[43]. However, it is worth noting that our study estimates the prevalence in the general population rather than solely at birth. Although our estimates draw on routine clinical data from Eastern Denmark (approximately 2.75 million inhabitants), this population is considered broadly representative of Denmark (approximately 6 million people), supporting extrapolation of our findings to the national level. Denmark, is, however, characteristic of a WEIRD (Western, Educated, Industrialised, Rich and Democratic) population [50], and caution is therefore warranted when generalising to other countries. Despite these limitations, our MCD prevalence estimates may help support and motivate future population-based studies and promote the adoption of similar methodological approaches.

## Conclusion

In this study, we provided the first population-based estimates of MCD prevalence associated with epilepsy in a large cohort and showed that epilepsy prevalence in the general population is consistent with that reported in the literature for similar populations. Characterising the epidemiology of each type of epilepsy-associated MCDs can better inform the increasingly diverse treatment modalities, particularly when it comes to surgical interventions. To date, population-wide prevalence estimates for many MCDs have been missing. Our study contributes to a better understanding of the frequencies of multiple of these abnormalities in patients with epilepsy. This can help both clinicians and researchers allocate resources to improve the outcomes of affected individuals. Future efforts should focus on harmonising the estimates for each type of brain structural abnormality associated with epilepsy across existent and future literature.

## Acknowledgments

This work was supported by the Lundbeck Foundation BrainDrugs grant (R279–2018–1145). JDiC received funding from the Lundbeck Foundation (grant R389-2021-1596) through Neuroscience Academy Denmark.

## Author contributions

The study was conceptualised by JDiC, LC, and MG. JDiC performed the formal analysis, and LC, VB, and AS the data analysis. HK and MP performed data annotation. The original draft writing was performed by JDiC, LC, and MG. All authors reviewed and approved the final version of the manuscript.

## Ethical Publication Statement

Ethics approval for data collection was granted by the Danish National Ethics Committee (National Videnskabsetisk Komité, Denmark, protocol number CVK-2112460), and approval for data processing was granted by the Data Protection Authorities of Denmark.

## Data availability statement

The data supporting the findings of this study contain personal information and therefore cannot be made publicly available.

